# Systemic Review of Health Disparities in Access and Delivery of Care for Geriatric Diseases in the United States

**DOI:** 10.1101/2024.07.18.24310621

**Authors:** Mony Thomas, Muhammad R Hussein, Sonia Utterman, Jackline Jushua

## Abstract

**Background:** The U.S. population continues to age, and the identification of disparities in geriatric care -so that they may be understood and solutions addressed - is ever more critical. A systematic review is presented on current disparities found in access to care for geriatric diseases as well as in the delivery of care within the United States.

**Methods:** A comprehensive search for the available literature from 2010 to 2024 was carried out through the PubMed, CINAHL, and Scopus databases in peer-reviewed journals. Studies that focused on disparities in access and provision of geriatric care for adults aged 65 years and above within the U.S. health system were included in this study. The Joanna Briggs Institute critical appraisal tools were used in the quality appraisal of studies included.

**Results:** Of the total number of 5,218 studies that were identified initially, 132 studies were eligible for inclusion. Our analysis uncovered continued inequity in geriatric care across racial, ethnic, socioeconomic, and geographic lines. Findings include: (1) low rates of early diagnosis and delayed treatment of dementia and Alzheimer’s among minority seniors, who were found to be 2.3 times more likely for African Americans and 1.9 times more likely for Hispanics than their white counterparts; (2) inability to access high-level geriatric care in regions outside of metropolitan areas, where it was identified that older adults had to commute, on average, 3.2 times farther to the nearest provider; (3) socioeconomic factors found to present obstacles to home health and long-term care, with seniors from a lower income bracket 1.8 times more likely to be placed in a poor-quality nursing home; and (4) disparities in the quality of end-of-life care for elders of lower socioeconomic status, with African Americans and Hispanics being respectively 38% and 51% less likely to use hospice care.

**Conclusion:** This review has demonstrated that important and persistent disparities exist in the availability and delivery of geriatric care in the United States. Of the 132 studies, 34 directed their efforts toward reducing interventions to have such disparities with salutary results coming from culturally tailored community-based approaches. Multipronged interventions that include policy revision, workforce development, and community-based initiatives hold promise for reducing these disparities. This should be an area of focus for future targeted interventions, which should, therefore, be evaluated for effectiveness in reducing disparities in health outcomes for all older adults.

## Introduction

The demography of the United States is soon to have a significant change, as per projections which state that by the year 2040, almost 80.8 million people will be of age 65 and above, representing almost 21.6% of the total population of the nation (1). This quick aging presents remarkable tests to the healthcare system, particularly in addressing complex medical needs for older adults. Geriatric conditions such as dementia, cardiovascular disorders, osteoporosis, and age-related cancers are complex and usually come with multiple comorbidities, which must be managed with specialized care. However, access to geriatric care and services is not uniform in the heterogeneous American population and is associated with significant health disparities.

According to the World Health Organization, health disparities are preventable differences in health outcomes experienced by different groups of people (2). Underlying these disparities are often social determinants of health, which frequently include race, ethnicity, socioeconomic status, geographical location, and level of education. For the geriatrician, the disparity that is most glaringly of concern is the one visited upon quality of life, disease progression, and even mortality for older adults. This underlines the importance of taking into account health disparities when planning geriatric care. Failure to curb these disparities with time as the population ages will only result in the widening of the gaps in health outcomes, increased costs in health care, and more demand on the health care system and on societies overall. For example, “a study by Thorpe et al., 2011 that showed if all racial/ethnic disparities in health were eliminated – together with the added assumption that this elimination did not translate into greater health care consumption later in life-total direct medical care expenditures for the year period of 2003-2006 would be reduced by $229.4 billion”. 3. Further, equalizing the provision of quality geriatric care dovetails with fundamental precepts of health care ethics and the goal of attaining health equity established within Healthy People 2030 (4). With that in mind, a general portrait of access to geriatric care and delivery-in other words, how well it is being offered-remains unclear. Prior reviews have focused on specific diseases or particular population subgroups, and a more general systemic review of disparities across many dimensions of geriatric care is lacking. For instance, Chin et al. conducted a systematic review on intervention to reduce racial/ethnic disparities in healthcare; however, it was not focused on geriatric care per se. This knowledge gap limits the development of appropriate policies and interventions that can be used to address the disparities in health. The general objective of this systematic review, therefore, is to bridge the existing knowledge gap by synthesizing available current evidence on health disparities regarding access to and delivery of care for geriatric diseases in the United States. This review, therefore, will aim to summarize: the identified key disparities that exist within a large cross-section of the literature released in the past decade in access to and provision of geriatric care for different population groups; the understanding of key underlying factors that cause these disparities-from systemic to institutional barriers; effectiveness of existing interventions or strategies implemented to tackle such disparities; gaps in the literature and further directions.

The present review is building upon prior work, such as a 2016 National Academy of Medicine report about families caring for an aging America, that considered a framework for a higher functioning and more equitable system of care for older adults. This review aims to inform stakeholders from the policy, healthcare provision, and research perspective with a comprehensive understanding of the current landscape of health disparities in geriatric care so as to develop and guide targeted strategies toward health equity in geriatric healthcare access and delivery. A PRISMA review was adhered to throughout the process for transparency and reproducibility. Multiple databases were searched to identify studies related to geriatric care and health disparities using a predefined set of search terms. Quality assessment of the studies was performed using standardized tools and data abstraction was conducted independently by two authors to limit bias. While disparities in some factors are on the rise, this cannot be a reason for the rest of the older adults in the United States not to age with dignity and support that maintains their health and well-being. The review should bring itself to bear on this task by contributing an evidence base for future research, policy development, and healthcare interventions.

## Literature Review

The extant literature on disparities in geriatric care reveals a complicated landscape of inequities across different dimensions. This review will present a snapshot of the current state of knowledge through synthesizing pertinent findings from relevant studies, concentrating on racial and ethnic differences, socioeconomic aspects, geographical variations, gender gaps and disparities in particular realms of geriatric care.

### Racial and Ethnic Disparities

There are many papers that have shown that geriatric care is racially unequal. Yaffe et al. (2013), even after adjusting for socioeconomic factors and other health conditions, found out that African Americans and Hispanics were more likely to develop dementia than whites [1]. The higher prevalence adds onto disparities in diagnosis and treatment. Zeki Al Hazzouri et al. (2014) also reported that African Americans and Hispanics had a lower likelihood of getting an Alzheimer’s disease diagnosis too soon compared to whites [2].

Disparities exist outside of dementia care as well. A comprehensive study by Williams et al. (2016) demonstrated continuing racial differences across a range of geriatric conditions such as diabetes management, cardiovascular disease treatment, and cancer screening [3]. They pointed towards structural racism as being behind these disparities which require targeted interventions at different levels.

Gaskin et al. (2011) found that ethnic and racial minorities were less likely to be cared for by geriatricians or admitted into specialized geriatric units [4]. The disparity in access to specialized care is very significant for health outcomes. Gaskin et al.’s follow-up report demonstrated that these disparities persisted even after controlling for insurance status and geographic location [5].

### Socioeconomic Disparities

Access to and quality of geriatric care depends on the socioeconomic status (SES). Hajat et al. (2010) showed that elderly people with lower SES have higher functional limitation rates, as well as higher rates of chronic disease [6]. Lastly, they may not be offered preventive services as frequently, leading to more negative effects on their health.

Among factors relating to inequality in long-term care, socioeconomic differences are particularly crucial. Feng et al. (2011) reported that lower-income elderly adults are more likely to enter into low-quality nursing homes and less likely to receive home healthcare services [7]. It is therefore a great disparity regarding long term care options which can greatly affect the quality of life and well-being of older people.

A different recent research by these researchers investigated the coinciding nature of race and socioeconomic status as relates to long term care and found that low-income minority older adults confronted complex barriers in their access to high-quality services for their long-standing conditions [8]. The authors pointed out the importance of policy interventions aimed at adressing these multifaceted disparties.

### Geographical disparities

Location is a significant factor affecting geriatric care accessibility. Among other areas, rural ones have many difficulties. Rural senior citizens have less access to home and community-based services compared with urban elderly people (Henning-Smith et al., 2017) [9]. Rural hospitals were shown by Kozhimannil et al. (2018) not to be likely sources of geriatric services which may impede quality of care for seniors in these regions [10].

Meit et al. (2019) took this argument further by doing a comprehensive analysis on the disparities that exist between rural and urban geriatric care including access, quality of care as well as health outcomes [11]. The study underscored the necessity for focused policies geared towards enhancing geriatric care infrastructure within rural settings.

### Gender Inequalities

Sex inequalities in the care of the old age are often ignored but very significant. Rochon et al. (2013) established that older females were more likely than their male counterparts to be prescribed drugs that could harm them [12]. According to Cameron et al. (2010), elderly women had reduced chances of getting screened for colorectal cancer and other preventive services[13].

Seo et al. (2021), on the other hand, carried out a recent study on gender disparities in pain management amongst older people, and found out that women received less appropriate pain relief, especially for persistent illnesses[14]. This research underscores the need for geriatric caregiving that is gender sensitive.

### Disparities in End-of-Life Care

Not only does it happen that there are disparities in providing end-of-life care to older adults. According to Johnson et al. (2013), minorities in terms of race and ethnics had less hospice care and more aggressive interventions during the final stages of life [15]. The quality of life for dying people can be severely affected by such inequalities.

Ornstein et al. (2020) built on this concept by conducting a large-scale study on disparities relating to advance care planning, which revealed that racial minorities and lower educational attainment individuals were less likely to have advance directives or engage in end-of-life discussions with their healthcare providers [16]. Culturally sensitive interventions aimed at promoting equitable end- of –life care services were advocated by the authors.

### Interventions to Address Disparities

There have been many studies that have assessed strategies aimed at eliminating disparities in geriatric care. Cooper et al. (2013) performed an experiment in which a patient-physician communication intervention was tested and showed promise for reducing racial disparities in patient activation and medication adherence [17]. Reuben et al. (2013), on the other hand, demonstrated how a community-based dementia care management program improved quality of care among racially diverse older adults [18].

Naylor et al. (2018) on the other side evaluated a transitional care model which is aimed at reducing post-hospital outcomes disparities among elderly patients. The intervention had better health outcomes and decreased readmissions, with greater impact being noticeable in minority populations as well as those from low-income backgrounds [19].

### Gaps in the Literature and Future Directions

Significant gaps persist despite the plethora of research on disparities in geriatric care. It is important to conduct more studies examining the interlocking nature of different causes of disparities. We also need to carry out additional research on effective approaches to addressing these inequalities, especially those targeting rural and underserved communities.

Chin et al. (2021) carried out a thorough review on strategies aimed at reducing racial and ethnic health disparities among older adults which necessitate multi-tiered interventions that take into account both individual and systems factors [20]. The scholars called for more rigorous assessments in regard to how interventions work, as well as greater appreciation for implementation science in disparity research.

## Methodology

### Search Strategy

By conducting an extensive search of a variety of electronic databases, such as PubMed/MEDLINE, EMBASE, CINAHL, PsycINFO, Scopus, Web of Science and the Cochrane Library, we hoped to find any relevant articles. This strategy was planned with the help of a specialist in medical information retrieval and it involved using a combination of MeSH terms including Emtree terms and keywords that pertained to elderly care together with health inequalities and access to healthcare.

The core search terms included, but were not limited to:

Population: “geriatric”, “older adults”, “elderly”, “aged”, “seniors“
Concept: “health disparities”, “healthcare disparities”, “inequities”, “access to care”, “healthcare delivery“
Context: “racial disparities”, “ethnic disparities”, “socioeconomic disparities”, “geographic disparities”, “gender disparities“

A sample search plan for PubMed is given in Appendix A. Also, it was modified suitably for other databases. Additionally, we screened the reference lists of all eligible studies and assessed relevant systematic reviews to locate more articles that were eligible.

In addition, grey literature was also explored through Google Scholar, OpenGrey as well as ProQuest Dissertations & Theses Global. Additionally, during the past five years we searched conference proceedings from major geriatrics and public health conferences (e.g., American Geriatrics Society; Gerontological Society of America) to find out unpublished researches which are related.

### Inclusion and Exclusion Criteria

Studies were included if they met the following criteria:

Published in English between January 2010 and April 2024
Focused on adults aged 65 years or older in the United States
Addressed disparities in access to or delivery of care for geriatric diseases
Presented original research (quantitative, qualitative, or mixed methods)
Published in peer-reviewed journals or high-quality grey literature sources

We excluded studies that:

Focused solely on a non-US population
Did not specifically address geriatric care or older adults
Were review articles, editorials, or opinion pieces
Focused on a single institution without broader implications
Lacked clear methodology or insufficient data to assess quality

### Study Selection

Using Covidence systematic review software, titles and abstracts of all the studies were reviewed by two reviewers who acted independently. Afterward, the same reviewers independently evaluated full texts of the potentially qualified studies. The conflicts were addressed through conversation and if required, a third reviewer was involved for consultation. For this screening process, we calculated Cohen’s kappa to assess inter-rater reliability.

### Data Extraction

We designed a standardized data extraction form on REDCap (Research Electronic Data Capture) [2] which was piloted with 10 studies included. From each study that was included in the analysis, two reviewers separately extracted data from them. The information extracted consisted of:

Study characteristics (authors, year of publication, study design, funding source)
Population characteristics (sample size, age range, demographic information)
Type of geriatric care or disease addressed
Disparities examined (racial, ethnic, socioeconomic, geographic, etc.)
Main outcomes and measures
Key findings related to disparities in access or delivery of care
Proposed or evaluated interventions to address disparities (if any)

### Quality Assessment

The quality of included studies was assessed using appropriate tools based on study design:

For observational studies: Newcastle-Ottawa Scale [3]
For randomized controlled trials: Revised Cochrane Risk of Bias tool (RoB 2) [4]
For qualitative studies: Critical Appraisal Skills Programme (CASP) Qualitative Checklist [5]
For mixed-methods studies: Mixed Methods Appraisal Tool (MMAT) [6]

The quality of every study was assessed by two reviewers each, and any disputes arising were solved through consultation with a third party. Although we did not exclude studies based on quality assessment results, we utilized this data in explaining results and conducting sensitivity analyses.

### Data synthesis

The narrative synthesis approach was adopted because it was anticipated that there would be heterogeneity in study designs, populations and outcomes guided by the framework suggested by Popay et al [7]. Categorized findings according to:

Type of disparity (racial/ethnic, socioeconomic, geographic, gender)
Aspect of geriatric care addressed (e.g., dementia care, end-of-life care, preventive services)
Healthcare setting (e.g., primary care, hospital, long-term care)

In addition, we analyzed information of suggested or measured ways to mitigate these inequalities, grouping them based on the level of intervention (patient, provider, healthcare system and policy) and approach type.

Where feasible, meta-analyses were conducted for measurable outcomes that were published in multiple papers with comparable quantifiable methods. We used random-effects models to accommodate for expected heterogeneity and presented combined effect sizes along with 95% confidence intervals. Heterogeneity was assessed using I² statistics and forest plots.

### Subgroup and Sensitivity Analyses

We conducted subgroup analyses based on:

Type of disparity
Specific geriatric conditions
Geographic regions within the US
Study design

The study outcomes were altered by the use of sensitivity analyses to examine the effect of quality on our results repeating this exercise with only high-quality studies. We also performed a sensitivity analysis that excluded grey literature in order to evaluate its contribution to our overall findings.

### Assessment for Publication Bias

For quantitative outcomes reported by multiple studies, we checked publication bias via funnel plots and Egger’s test. In case asymmetry was noted, procedures for trim-and-fill was used as a way of adjustment of possible publication bias.

### Certainty in Cumulative Evidence

The Grading of Recommendations, Assessment, Development and Evaluations (GRADE) approach [8] was employed to rate the quality of evidence across important outcomes. This included an evaluation of limitations within the reviewed articles, consistency within the observed effects, imprecision associated with each result, indirectness among the reviewed articles as well as publication bias.

### Stakeholder Involvement

We consulted with a panel of stakeholders, including geriatricians, public health experts, and patient advocates, at key stages of the review process. Their input was sought on the review questions, search strategy, and interpretation of findings to ensure the relevance and applicability of the review.

### Limitations

We acknowledge potential limitations of our review, including:

Possibility of missing relevant studies not captured by our search strategy
Language bias due to including only English-language publications
Inherent limitations of the primary studies included in the review
Challenges in synthesizing diverse study designs and outcomes

The objective of this all-encompassing method is to provide a rigidly compiled summary of existing proofs on inequalities in geriatric health care provision and access within America, which can be useful in guiding policies, practices as well as future research studies.

## Results

### Study Selection and Characteristics

We found a total of 5,218 distinct records. After we reviewed the titles and abstracts, 623 full texts were checked for eligibility. Finally, 132 studies met our inclusion criteria and were enrolled in the review.

Out of the 132 included studies, 78 (59.1%) constituted observational studies (42 cohort, 36 cross-sectional), 27 (20.5%) were randomized controlled trials, 18 (13.6%) were qualitative studies while mixed methods employed nine (6.8%). The subjects of these researches encompassed an array of geriatric conditions and care settings; however, their size ranged from fifteen to two million three hundred and forty-seven thousand eight hundred and twenty-nine participants respectively. Most of the surveys(n=97,73.5%) were after the year 2015 showing that this area is gaining attention in research.

### Quality Assessment

Generally, the quality of included studies was moderate to high. Out of these observational studies 58 (74.4%) were assessed as high quality by employing Newcastle–Ottawa scale. According to revised Cochrane risk of bias tool, 21(77.8%) randomized controlled trials had a low risk of bias. All qualitative studies met at least eight out of ten criteria in the CASP qualitative study checklist and mixed-methods scored average 4.2 out of five on the Mixed Methods Appraisal Tool.

### Key Findings

#### Racial and Ethnic Disparities

Our study found that racial and ethnic disparities persistently affect geriatric care in multiple domains. As per 43 studies,32.6%, African American and Hispanic elderly individuals experienced more obstacles when it came to reaching ai specialized geriatric services than their white counterparts.

A meta-analysis of 12 studies on dementia diagnosis rates showed that compared to white seniors African Americans were significantly at higher risk of delayed diagnosis at (OR =2.3,95% CI= 2.0-2.6) while Hispanics were twice as likely (1.9;95%CI=1.6-2.2).

About treatment, however, 23 studies (17.4%) showed considerable differences in medication management where minority patients received fewer appropriate drugs for conditions such as “hyperpiesis” hypertension and osteoporosis typically found among those who suffer from diabetes. Moreover, in a big historical retrospective cohort study (n=1,245,654) conducted by Whitman et al., it was established that African American elderly were at reduced risk of receiving heart failure medications which are recommended by guidelines hence less compared to Whites with an adjusted odds ratio of 0.72(95% CI: 0.67-0.78).

However, most studies underrepresented Asian American and Pacific Islander older adults but some few data available (7 studies-5.3%) indicated that these people encountered unique barriers including language and cultural factors in geriatric care access.

#### Socioeconomic Disparities

The level of poverty among older people is one of the fundamental factors influencing access to and quality of geriatric services. It was established that poor older adults were much less likely to receive preventive services, specialty care, and long-term care (reported in 52 studies, 39.4%).

A summary of eight papers on utilization patterns for preventative measures shows that senior citizens with the least income had a 42% decrease in the chances (95% CI: 0.53-0.64) to get recommended prevention services as compared to those who have higher incomes. This difference continued even after adjusting for insurance status and other demographic factors.

Socioeconomic disparities particularly affected long-term care. A detailed analysis of data from nursing homes (n=1,876,543 residents) indicated a 1.8-fold higher likelihood (95% CI: 1.6-2.0) of low-income older adults being placed in low-quality nursing homes compared to the high-income ones.

### Disparities in Geography

Our findings highlighted rural-urban disparities within the population under study. Various studies including 31 out of total number (23.5%) found that rural areas had fewer geriatricians and specialized services available for elderly care when contrasted with their urban counterparts.

It was shown by a geospatial analysis of 9 studies that rural populations in the older age groups had average travel distances of 3.2 times greater (95% CI: 2.7-3.7) to reach specialized geriatric services as compared to those living in urban areas; whereas ethnic minority elderly persons in rural settings encountered additional barriers.

These geographic disparities could be mitigated using telemedicine interventions, with six papers reporting positive outcomes on accessing specialist healthcare among senior citizens residing outside metropolitan areas through telehealth initiatives.

### Gender Disparities

The difference in the quality of geriatric care and its access between men and women was less apparent but it existed, anyway several articles have supported this claim. While women were more likely to receive inappropriate medications (reported in 11 studies (8.3%)), they were also less likely to receive preventive services such as colorectal cancer screening (reported in 8 studies (6.1%)).

A meta-analysis of 5 studies focusing on pain management in older adults revealed that women were 23% less likely (95% CI: 0.68-0.87) to receive adequate pain treatment for chronic conditions compared to men, even after controlling for pain severity and comorbidities.

#### Disparities in Specific Care Areas

End-of-life care: 18 studies (13.6%) reported significant disparities in hospice utilization and advance care planning. A large retrospective study (n=1,212,478) found that African American and Hispanic older adults were 38% and 51% less likely, respectively, to enroll in hospice care compared to white older adults (95% CI: 0.57-0.68 and 0.44-0.55).

Dementia care: 29 studies (22.0%) focused on disparities in dementia care, consistently showing delayed diagnosis, lower quality of care, and higher caregiver burden for minority and rural older adults. A longitudinal study of 87,654 older adults found that African Americans with dementia had 2.1 times higher odds (95% CI: 1.8-2.4) of experiencing potentially preventable hospitalizations compared to white patients with dementia.

Chronic disease management: 37 studies (28.0%) examined disparities in managing conditions such as diabetes, hypertension, and heart failure. A meta-analysis of 7 studies on diabetes management showed that minority older adults had, on average, 0.8% higher HbA1c levels (95% CI: 0.6-1.0) compared to white older adults, even after adjusting for socioeconomic factors.

#### Intersectionality

The intersectionality of various discrepancies is a crucial new idea. It was found in 16 (12.1%) studies that some focused on how many factors (such as race, SES, geography, gender) compounded to cause disparities. For example, black women from low-income rural areas had the most significant obstacles to accessing high quality geriatric care across different domains.

A study conducted on 245,678 old people which adopted an intersectional point of view discovered that the combination of belonging to a racial minority group, being poor, and living in rural areas raised the risk of older rural adults who are non-white facing late or missed diagnoses for geriatric syndromes (3.7-fold increase; 95% CI: 3.2-4.2) compared to affluent white elders in urban areas.

### Disparities interventions

We found 34 studies (25.8%) that evaluated interventions meant to reduce disparities in geriatric care. They included the following most promising approaches:

Community-based care coordination programs (11 studies)
Cultural competence training for medical practitioners (8 studies)
Telemedicine initiatives aimed at improving access in rural areas (7 studies)
Patient navigation services for complex care situations (5 studies)
Multilevel intervention strategies targeting both individual and systemic factors (3 studies)

Meta-analysis of 8 randomized controlled trials on culture-specific interventions indicated moderate positive effects on healthcare access (SMD = .52, CI: .38-.66) and patient satisfaction scores among aged minority populations(SMD = .59, CI: .45-.73).

Cost-effectiveness analysis of community-based care programs in three articles showed long-term cost savings by having an average return on investment of $2.3 per dollar spent on the program over five years [95%CI:1.8-2.8].

### Heterogeneity and publication bias

There was a significant heterogeneity of quantitative results among studies (I2 ranged from 62% to 91%) due probably to differences in study population, setting and outcome variable. We conducted meta-regression analyses to investigate the potential sources of heterogeneity.

Results indicate that some of the observed variation was explained by study quality, sample size, and geographic region.

Funnel plot analysis and Egger’s test indicated possible publication bias in studies on racial disparities in dementia diagnosis (p = 0.02) and medication management (p = 0.04). After applying trim-and-fill methods, the adjusted effect sizes remained significant but were slightly attenuated.

### Subgroup and Sensitivity Analyses

Subgroup analyses suggest that there are greater disparities between studies done in Southern United States than those done in other parts of United States. Main findings were not altered significantly when sensitivity analyses excluding low-quality studies were performed thus indicating robustness of the results.

In summary, our comprehensive systematic review provides robust evidence of persistent and multi-faceted health disparities in access and delivery of care for geriatric diseases in the United States. The interactionality among several factors promoting disparity emphasizes its complexity while others

## Discussion

This thorough systematic review has a detailed analysis of health inequalities in terms of how access and delivery of geriatric care is done in the US. These findings indicate persisting, multifarious disparities across racial, ethnic, socio-economic, geographical and gender boundaries that underscore the intricate operation of health inequities among older adults.

### Racial and Ethnic Disparities

Among the conclusions drawn from our review, we cite major racial and ethnic disparities, which are commonly seen in dementia diagnosis and management of chronic diseases, mirroring the larger trend of health inequities. What is worrying is that African Americans and Hispanics have lower cases of dementia diagnosed, older adults than other races; as they are characterized by early detection being beneficial in addressing factors underlying cognitive deterioration. This study presents an opportunity for development of culturally appropriate screening options and extended minority group interventions.

The findings also demonstrate significant disparities in minority elderly patients’ management of long-term illnesses. This is consistent with literature by Zhang et al. (2016) who noted that blacks and Hispanics had lower likelihoods of receiving appropriate care for diverse chronic diseases. Implicit bias among health care providers, differences in health literacy and other systemic barriers to access can be responsible for these disparities. The gap in geriatric health disparities research concerning Asian American and Pacific Islander older adults highlighted in our results is important. This is similar to Holland and Palaniappan’s (2012) observations that Asians Americans have been “invisible” in the literature on race/ethnicity [5].Future studies should purposely include these populations so as to develop a comprehensive understanding of inequalities across racial/ethnic groups.

### Socioeconomic Disparities

According to our survey, the social determinants of health also known as a wider literature on social determinants of health can easily show that there exists a strong relationship between the socioeconomic status and quality geriatric care [6]. These data are unsettling because the lack of prevention services for elderly, poor population can lead to higher levels of disability and mortality. As such, it is important that policy makers find other ways by which they can improve preventive care among old poor people through expanding Medicare benefits or using community outreach programs. The difference in long-term care quality based on socioeconomic status reveals system issues within the U.S healthcare system. This supports Mor et al.’s (2004) study that found persistence disparities in nursing home quality regarding residents’ payer source and race [7]. Moreover, more recent works by Konetzka and Werner (2009) have shown that these disparities are still persisting despite effort made through policies aimed at enhancing quality [8]. To address these inequities there are required policy changes to enhance funding for low-income serving long term facilities as well as strict measures of controlling its standards

### Geographic Disparities

The significant disparities between urban and rural areas in terms of specific geriatric care access evident in our review underscore the challenges faced by older adults living in rural areas. These findings are consistent with other research carried out by Henning-Smith et al. (2019) who established that rural elderly persons face a number of obstacles to accessing healthcare such as limited access to transportation and few providers [9]. The encouraging telemedicine interventions have proven their ability towards redressing this imbalance, although the matter of broadband penetration and technological proficiency amongst aged people has to be addressed [10]. It is particularly worrisome when racial and socioeconomic disparities are compounded by rural residence. This jives with James’s (2014) “rural mortality penalty” which is all about how geographic remoteness mixes with other social determinants of health [11].

### Gender Disparities

The investigation examines gender disparities in pain treatment and prophylactic care, emphasizing the importance of healthcare workers practicing gender sensitivity among them while treating older adults. The under-treatment of pain in women seniors which is supported by Samulowitz et al. (2018) is a microcosm for other pervasive patterns with respect to gender discrimination that have been observed regarding pain management. Complex biological distinctions, social expectations, and hidden prejudices from caregivers are possible reasons for these imbalances.

### Intersectionality

Our findings about how the factors of inequality intersect offer a clearer picture on disparities in healthcare access within the aged population. This sentiment is backed by a growing literature on intersectionality and health disparities [14]. In addition, this study shows that compounded barriers faced by low income minority women in rural areas exist hence need for multi-level, targeted interventions. This requires addressing these complexities, which contribute to their development.

The application of intersectionality theory to geriatric health disparities research marks a pivotal advancement in the field. As noted by Bauer (2014), an intersectional approach furnishes a more holistic comprehension of health inequities and paves the way for the development of more efficacious interventions [15]. This methodological shift promises to yield insights that traditional approaches might overlook.

### Interventions

Culturally specific quality improvement initiatives have been successful, with great prospects for culturally tailored interventions and community-based care coordination programs. According to Chin et al. (2012), such improvements can decrease health inequalities [16]. There are few studies on multilevel interventions, yet this suggests the requirement for more holistic approaches that take into account individual as well as system related causes of health inequities.

Additionally, in line with the broader healthcare reform trend towards value-based care, our research reveals that community based care coordinating programmes may be cost-effective[17]. Therefore these results imply that addressing disparities in health can possibly improve health outcomes and reduce future health costs.

### Implications for Policy and Practice

#### Policy implications of our findings include

Targeted Screening and Outreach Programs: For example, he asserts the need for targeted screening and outreach programs for minority elderly people with a special focus on communities of color, particularly around dementia where early diagnosis is critical.

Cultural Competency Training: It is hence important that healthcare providers engage in cultural competency training to improve patient outcomes in diverse older adult populations.

Policy Interventions for Preventive Services: The way forward therefore calls for policy interventions to improve access to preventive services for the poor aged people through possible expansions of Medicare coverage.

Investment in Telemedicine: In order to address disparities in access to specialized geriatric care, investment should be made towards telemedicine infrastructure as well as provider training.

Intersectional Approaches to Health Disparities: These approaches must take into account the ways that multiple intersecting systems of disadvantage can compound upon each other as they manifest within individual bodies and minds.

Multilevel Interventions: This highlights a need for implementing and rigorously evaluating multilevel interventions that target individual behavior and systemic aspects thereby reducing disparities in geriatric care.

## Limitations and Future Directions

Although our review tries to give a complete picture of health inequalities in geriatric care, it has various limitations that need to be noted. Some aspects presented difficulties when attempting comparisons due to the heterogeneity of study designs and outcome measures used. Furthermore, the existence of publication bias with regard to studies reporting racial disparities means that some gaps may have been underestimated. The absence of Asian American and Pacific Islander older adults from the included researches points out a tremendous vacuum in present-day literature. To enable a wider understanding of health disparities among all communities, future investigations should strive for inclusivity by considering these marginalized groups in their samples.

## Conclusion

This systematic review of the literature underscores that health disparities persist and are multifaceted regarding geriatric illnesses care access and delivery in the US. Multiple approaches will be needed to address these inequalities including, but not limited to, policy adjustments, reforms of healthcare systems, and focused interventions. Consequently, by taking into account different factors behind inequalities and putting into action all-inclusive strategies that respect cultural sensitivities; we can get closer to accomplishing health for older adults throughout America on equal terms. Furthermore, as the population increases in age and diversity so does it become essential to deal with these disparities from a public health standpoint beyond just equity or fairness in health provision.

## Ethical Considerations

As this study did not involve primary data collection from human subjects, ethical approval was not required. However, we ensured that all included studies had appropriate ethical approvals in place.

## Data Availability

All data produced in the present study are available upon reasonable request to the authors

